# The Accuracy of Cardiac Surface Conduction Velocity Measurements

**DOI:** 10.1101/2024.01.26.24301849

**Authors:** Edward J. Vigmond, Caroline Roney, Jason Bayer, Kumaraswamy Nanthakumar

## Abstract

**Background:** Conduction velocity (CV) is a measure of the health of myocardial tissue. It can be measured by taking differences in activation times from intracardiac electrodes. Several factors introduce error into the measurement, amongst which, ignoring the three dimensional aspect is a major detriment. The purpose of this paper was to determine if, nonetheless, there was a specific region where CV could be accurately measured. **Methods:** Computer simulations of three dimensional ventricles with a realistic His-Purkinje system were performed. Ventricles also included a dense scar or diffuse fibrosis. **Results:** A finer spatial sampling produced better agreement with true CV. Using a error limit of 10 cm/s as a threshold, measurements taken within a region less than 2 cm from the pacing site proved to be accurate. Error increased abruptly beyond this distance. The Purkinje system and tissue fibre orientation played equally major roles in leading to a surface CV which was not reflective of the CV propagation through the tissue. **Conclusions:** Surface CV measurements close to the pacing site taken with an electrode spacing of at most 1 mm, give reasonable estimates of tissue CV.

## 1 Introduction

Conduction velocity (CV) is an important measure to assess the health of cardiac tissue. It is sensitive to both structural changes, as well as ionic modifications. Structural changes, such as fibrosis, can lead to a more tortuous propagation which, in the absence of gap junctional remodelling, results in a slower macroscopic CV. Disruptions to gap junctions also leads to direct CV changes on the microscopic scale. Perturbations to sodium channels also directly influence CV, and ionic channels which modify the resting level also have an effect by modulating the level of inactivation of sodium channels. Thus, while CV changes are nonspecific, further elucidation is possible by considering the rate of change of voltage during the upstroke, and the degree of electrogram fractionation [1, 2].

Cardiac tissue is three dimensional which necessitates three dimensional recordings for accurate measurement. However, clinically measurements are restricted to the surface, whether it be by electroanatomic mapping systems or ECGi. Clinical systems are being developed which measure local, surface CV. These systems are evolving to be multipolar with small interelectrode spacing, allowing more accurate determination of activation from any direction. We have previously shown the inaccuracy of surface measurements when trying to localize intramural ectopic foci, as well as the error in computing CV [3,4]. Thus, the interpretation of these measurements should be treated cautiously, as there validity or utility need further clarification.

In this paper, we consider whether there are conditions under which recorded CV may be considered free from artifact and reflects the true CV, allowing it to be a diagnostic measure. We perform computer simulations — the only method being able to easily provide high density 3D CV— in a human geometry with a conduction system. Pacing is performed from different sites under different conditions, and the endocardially measured CVs, at different resolutions, are compared to the true CV.

## 2 Methods

### 2.1 Geometry

A three dimensional, human, finite element mesh was used with 450 micrometer discretization, yielding 5 million nodes and 26.4 million tetrahedral elements. A Purkinje system (PS) was added based on known anatomical detail, having a His bundle which divided into three fascicles in the LV, and a single fascicle to the RV bifurcating to form the moderator band and an apical extension [5]. See Fig. 1. There were 522 Purkinje-myocyte junctions through which electrical activity propagated between the myocardium and the Purkinje network.

**Figure 1.**
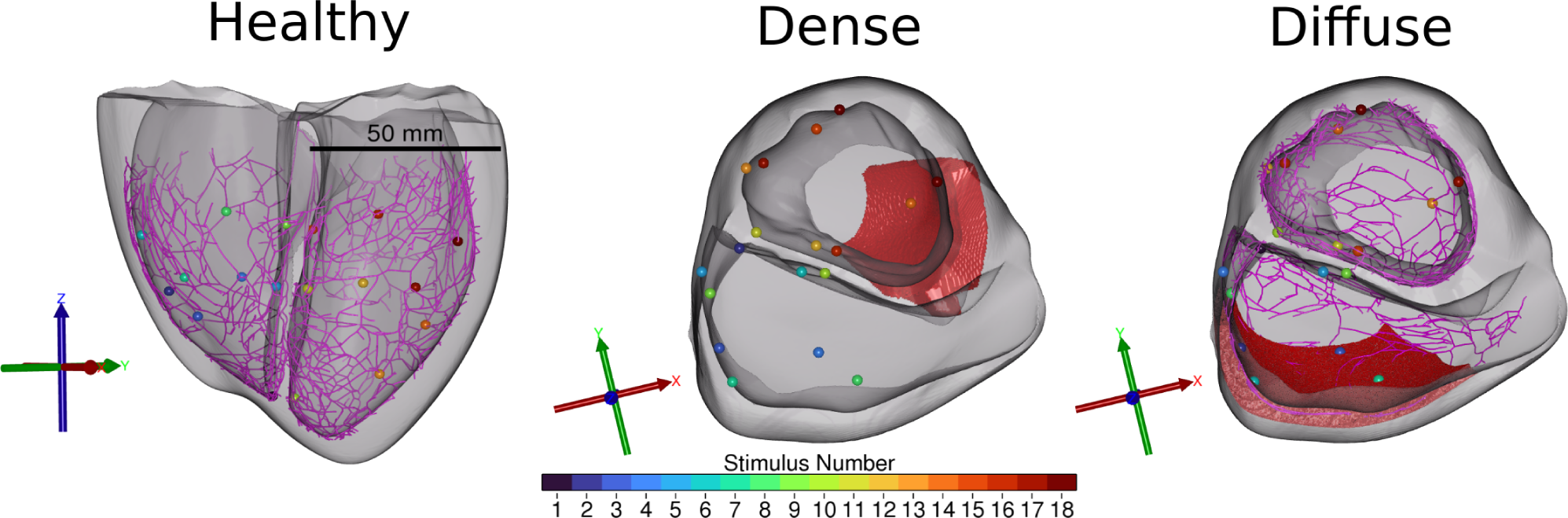
Experimental set up. Stimulation points are shown on the different views of the ventricles. Left: The Healthy heart had no fibrosis. The Purkinje system is depicted by the purple lines, and a scale bar is shown. Middle: the dense scar region is in red, and the Purkinje system is not shown. Right: The diffuse fibrosis is shown with the Purkinje system. Axes give the orientations of the hearts. The first 9 sites are in the RV and the rest in the left. Within each ventricle, numbering increases with distance from the base.

A dense scar was synthetically added to the anterior LV wall (Fig. 1 middle) by defining it in universal ventricular coordinates: 0.3 *< z <* 0.7, *π/*4 *< ϕ <* 3*π/*4, 0.1 *< ρ <* 0.9, *ν* = *−*1 [6]. The scar was not fully transmural, allowing surviving epicardial and endocardial layers. Within the dense scar, all tissue was considered dead and therefore not included in electrophysiological calculations. A region of diffuse fibrosis was also defined (Fig. 1 right) by universal ventricular coordinates and added to the RV free wall: 0.3 *< z <* 0.7, *−*3*π/*8 *< ϕ <* 0, *ρ ≤* 1, *ν* = 1. Within this region, CV was reduced by a factor of four in all directions. The PS was assumed to be unaffected by fibrosis and scar [7, 8].

### 2.2 Electrophysiology

The ten Tusscher ionic model for human ventricular cells was used for the working myocardium, and the Steward canine Purkinje ionic model was used for the PS. All simulations were performed with the carpentry electrophysiological simulator [9] using a time step of 25 *µ*s. Conductivities in the model were set to produce conduction velocities in the myocardium of 71.4, 21.7, and 12.9 cm/s in the longitudinal, transverse and sheet normal directions in accordance with values and ratios seen physiologically [10, 11]. Sinus activation of the heart as measured from simulated lead potentials took 134 ms.

A total of 18 endocardial pacing sites were used, evenly divided between the ventricle, spread along the major axis, i.e., *z*, with universal ordinates of 0, 0.5, and 0.7. See Fig. 1. A 2 ms long transmembrane current of 70 *µA/cm*^2^ was applied to a 1 mm^3^ volume at each pacing point, affecting only myocardium.

### 2.3 Conduction Velocity

Activation time was based on transmembrane voltage, taken as the instant when the transmembrane voltage exceeded −50 mV with a dV/dt*>* 5 V/s calculated with a resolution of 100 *µ*s. CV was calculated as the magnitude of the inverse of the gradient of activation times over each finite element. To arrive at different sampling resolutions, the endocardial meshes were extracted from the original fine mesh, and then resampled at edge lengths of 0.5, 1, 1.5, 2, 3, and 4 mm. Activation times were transferred to the coarser mesh by inverse distance weighting radial basis functions, and CV computed from these times on the coarser surface-only mesh. CVs were subsequently smoothed using Gaussian radial basis function weighting with a 4 mm radius. The gold standard was the CV measurements made on the fine mesh taking into account the 3D propagation. Errors reported are the difference between CVs derived from surface calculation versus those obtained from the full 3D propagation. Distances from pacing sites refers to the distances traveled through the myocardium to reach a recording site from the pacing point, determined by using an Eikonal solution with unit isotropic conduction velocity.

## 3 Results

### 3.1 Nominal case

The magnitude of the CV error is shown as a function of distance from the pacing site for all stimuli in Fig. 2. All curves have the same general shape. Very close to the pacing site, the error decreases from an initial value. After several millimeters, the error increases slowly. At some distance from each pacing site, when the ensuing wavefront started to cross the septum or reached an edge, the error rose rapidly to a high level and then returned just as rapidly to a stable level, as the activation once again propagated through the free wall. The large error was associated with breakthroughs when the wavefront simultaneously activated large regions of the contralateral septal wall.

**Figure 2.**
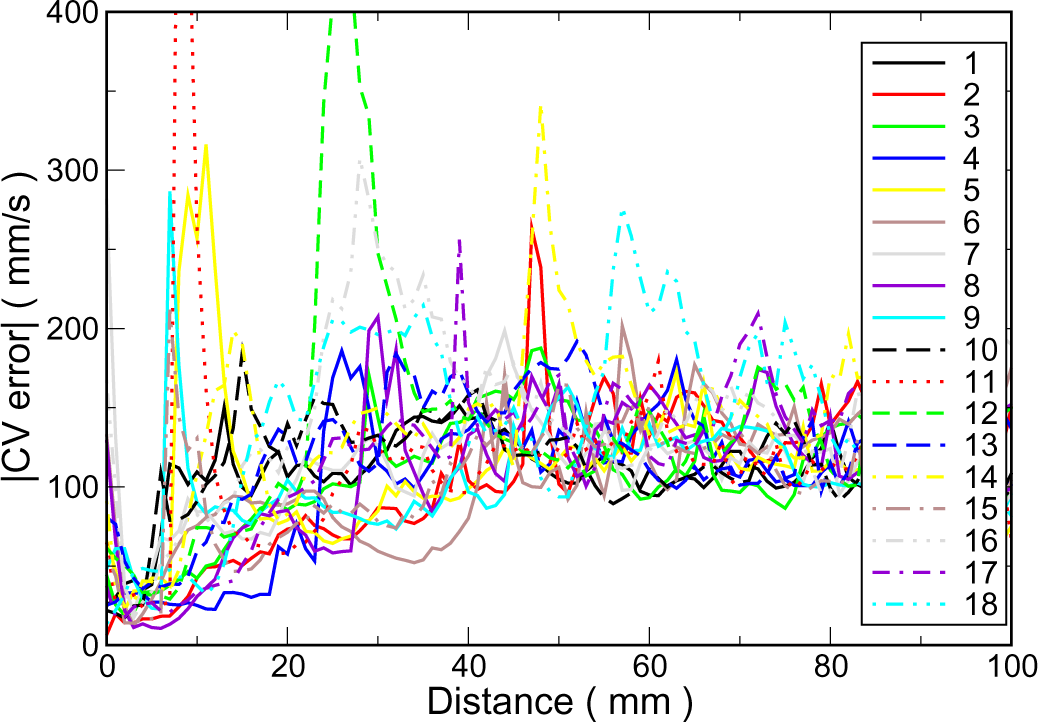
Magnitude of CV error as a function of distance from each pacing site. RV sites have solid curves while LV related curves are dashed. The grid resolution was 0.5 mm.

The case of pacing site #12, a mid-height anterior site, is shown in detail in Fig. 3. The error distribution is typical for all pacing sites. A low error region surrounds the pacing site with a larger error band running through it, which coincides with the endocardial fibre direction. The error transition is rather abrupt after 1–2 cm, depending on the direction travelled. The large error can be seen on the RV septum in the posterior view (Fig. 3 upper left). CV is consistently overestimated, and is only underestimated at the basal border where propagation hit a barrier.

**Figure 3.**
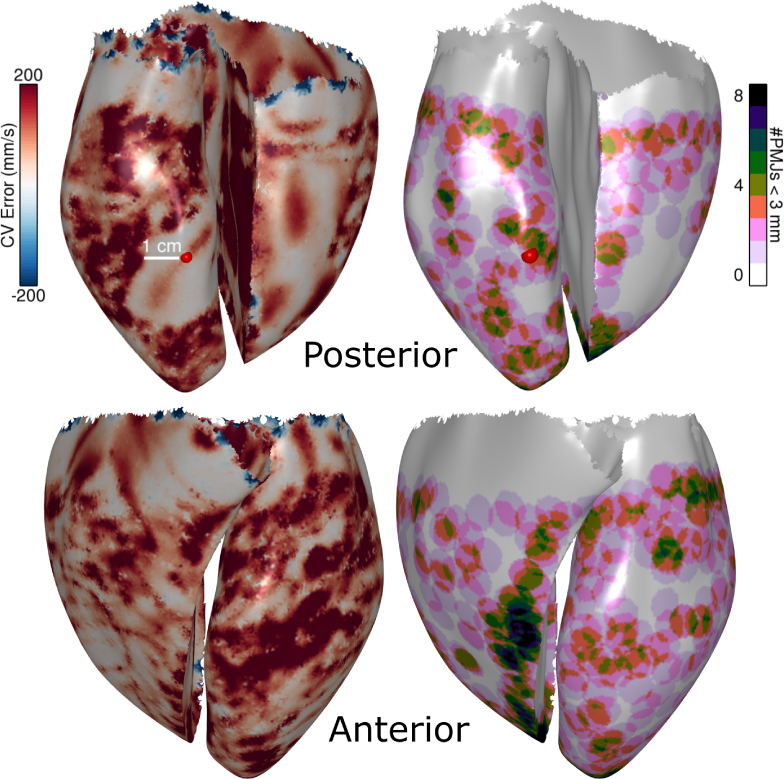
CV error and PMJ density. The pacing site #12 is indicated by the red dot. **Left**: The magnitude of the conduction velocity (CV) error on the endocardial surfaces at a resolution of 1 mm. A white scale bar indicates a distance of 1 cm. **Right**: The number of PMJs within a radius of 3 mm from each surface point is mapped.

The density of Purkinje-myocardial junctions (PMJs) is shown in the right panel of Fig. 3. There appears to be a correlation between junction density and error. Regions of high junctional density with large error are found (anterior mid LV, or high anterior LV), as are low junctional density regions with low error (basal regions, and lower posterior RV free wall). However, regions do not perfectly overlap as there are high junctional density regions with low error (anterior RV near septum), and low junctional density regions with high error (anterior RV base).

### 3.2 Effect of resolution

The error averaged over all pacing sites for each resolution is shown in Fig. 4. All resolutions showed a similar shape. Close to the pacing electrode, the error is low and reaches a minimum at a few millimeters away, followed by a steep rise with an abrupt peak, beyond which the error gradually changes. Beyond 10 cm (not shown), errors become quite large as errors are measured in the non-stimulated ventricle, and there are septal breakthroughs and wavefront collisions. The error was similar for the highest two resolutions, 0.5 and 1 mm edge lengths. For the other resolutions, the error was unacceptable.

**Figure 4.**
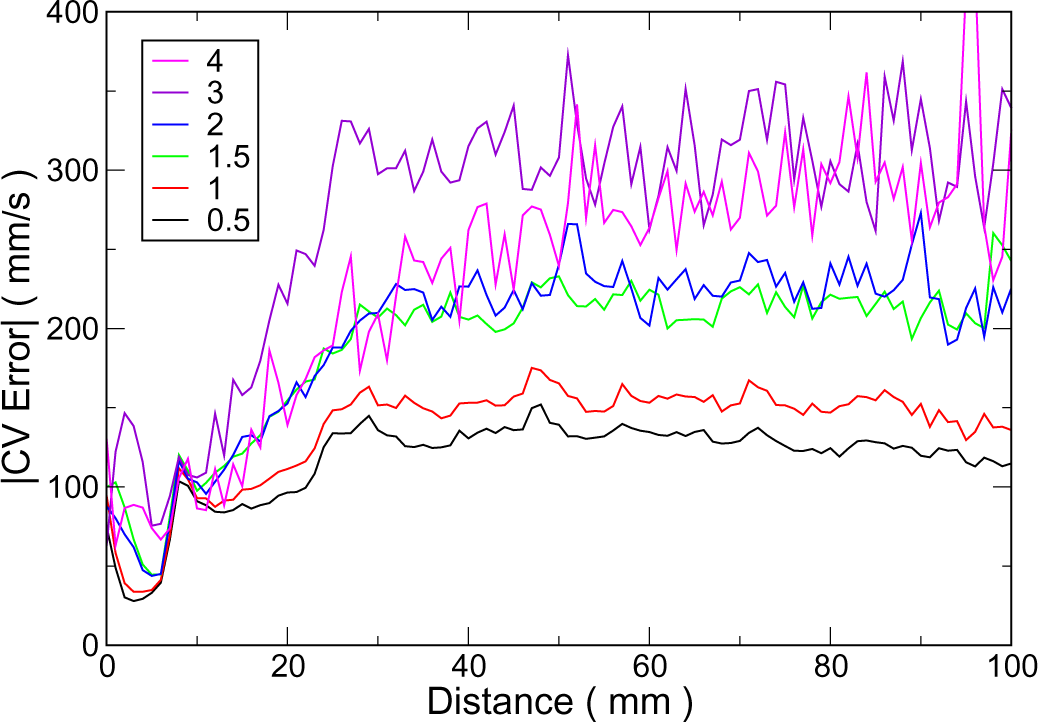
CV error versus distance from pacing site depends on mesh resolution. The average magnitude of the error in conduction velocity is plotted as a function of surface distance from all pacing sites for various mesh resolutions. Resolutions are indicated as average element edge length in mm.

### 3.3 Recording location

The CV error was analyzed for the endocardial regions divided into the free wall and septum for each ventricle. The magnitude of the CV error in each region was averaged over the region for all pacing sites in a particular ventricle. Results are summarized in Table 1.

**Table 1.**
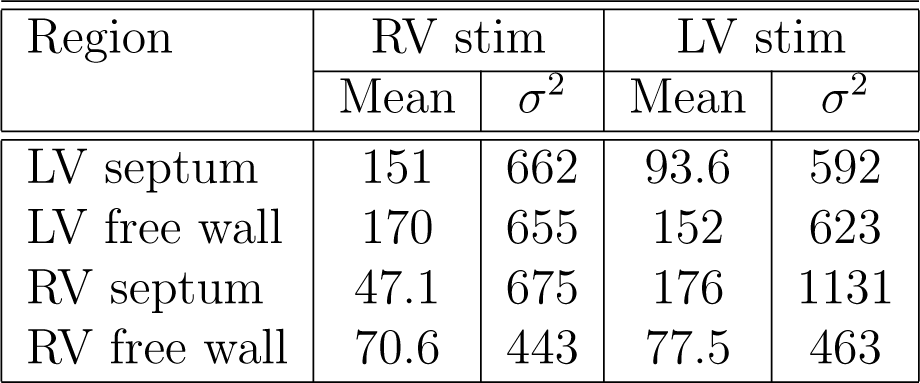
Average CV error magnitude (mm/s) and its standard deviation (mm/s) for each region as a function of the ventricle paced. Errors are averaged over all pacing sites in each ventricle for healthy ventricles. The grid resolution was 1 mm.

| Region | RV stim |  | LV stim |  |
| --- | --- | --- | --- | --- |
| | Mean | $\sigma^2$ | Mean | $\sigma^2$ |
| LV septum | 151 | 662 | 93.6 | 592 |
| LV free wall | 170 | 655 | 152 | 623 |
| RV septum | 47.1 | 675 | 176 | 1131 |
| RV free wall | 70.6 | 443 | 77.5 | 463 |

In all cases, contralateral measurements had higher errors than same cavity measurements. The LV free wall had the largest error regardless of ventricle activated. The RV free wall had the lowest overall error, while the RV septum was fairly accurate for RV pacing sites but very inaccurate for LV pacing.

### 3.4 Effect of Fibrosis

The effects of including a dense intramural scar, or diffuse fibrosis, are shown for a 1 mm resolution in Fig. 5. The error was different in very close proximity to the pacing, being far less for the healthy case and diffuse scar, but there was little difference farther than 1 cm. The scar region was not identifiable by CV changes over the scar, regardless of pacing site.

**Figure 5.**
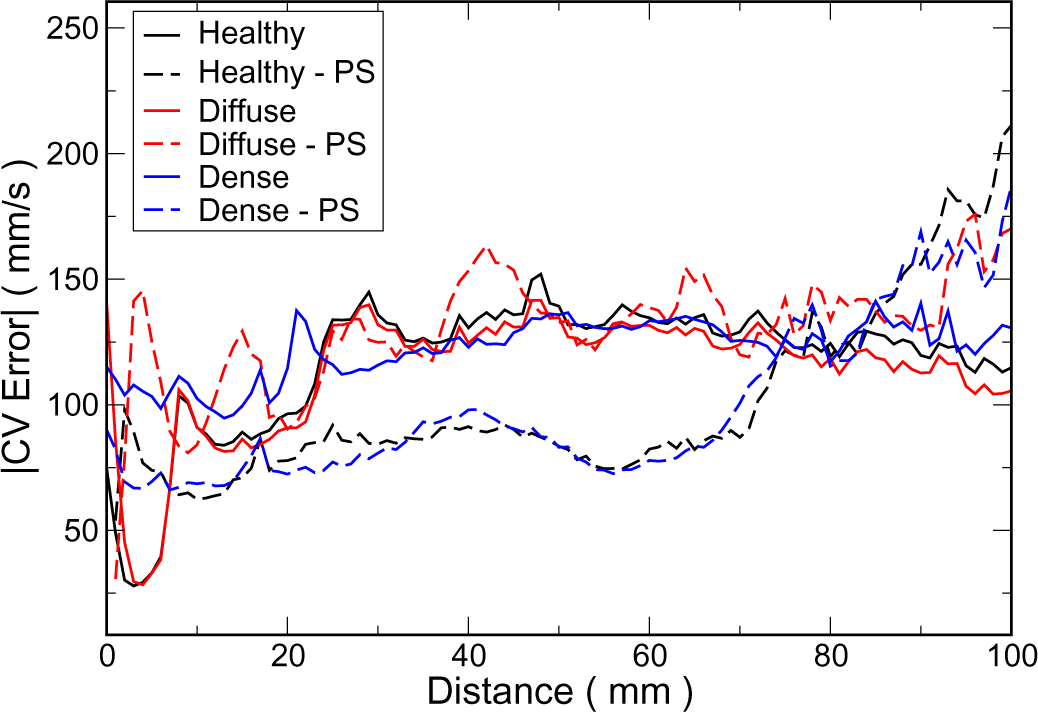
Effect of the fibrosis and Purkinje system on CV error. The magnitude of the error in conduction velocity is plotted as a function of surface distance from pacing site averaged over all pacing sites for 1 mm resolution. The error is shown for healthy ventricles (black), with diffuse fibrosis (red) and dense fibrosis (blue) with and without a Purkinje system (-PS). The resolution used was 1 mm.

The CV measured and the error are shown for pacing site #4, a lower anterior site in the RV, in Fig; 6. The region of diffuse scar is clearly seen in the CV map as a region of reduced CV. Very high CVs are seen on septal breakthroughs in the nonstimulated ventricle (bright yellow on the left). The CV error within 2 cm of the pacing site was very low, and the CV error over the fibrosis was relatively low, mostly less than 50 mm/s, but some high errors are also seen. Low CV was consistently seen over the diffuse fibrosis, regardless of pacing site, even contralateral. The activation pattern shows dense spacing of isochrones over the fibrosis with many breakthroughs (Fig; 6 left). These are more apparent in the fibrosis region since the PS was assumed to be unaffected and the lower CV of the myocardium did not allow enough time for wavefronts travelling through the tissue reach PMJs before activity emerged. The CV error in the LV was much higher and much less reliable.

### 3.5 Purkinje activation

The effect of the Purkinje system was ascertained by performing simulations with no Purkinje system (Fig. 5). As noted previously, there was little difference in error for the three cases with a Purkinje system present. For healthy tissue and dense scar, removing the Purkinje system decreased the error substantially, by approximately 50 mm/s, for distances between 1 and 7 cm. Diffuse fibrosis was little affected in this range. Close to the pacing site, diffuse fibrosis and healthy tissue errors were increased while dense scar CV error was decreased.

### 3.6 Factors Affecting Accuracy

The sensitivity of the CV to the relative speed of propagation was performed. The conductivities were doubled to achieve approximate increases of 1.41 fold. Sinus activation was affected as expected (see Fig. 7). Increasing the Purkinje CV decreased the total activation time and led to more pronounced breakthroughs and transmural propagation was allowed less time to propagate from neighboring previously activated junctions. Increasing the myocardial propagation speed did not decrease total activation time very much as apex to base conduction is primarily dictated by the Purkinje system except for the most basal portion. Increasing myocardial conduction velocity provides more interaction between basally oriented wavefronts and junctional activations that masked the breakthroughs.

Furthermore, the effect of fibre structure and the Purkinje system were also compared. Removing the Purkinje system had the same effect on accuracy as ignoring the fibre structure. When ignoring the Purkinje system as well as the fibre structure, the error was greatly reduced even at farther distances from the pacing site.

The effect of the relative changes in conduction velocity on the CV error is shown in Fig. 7 bottom. With a fast Purkinje system, the error in close proximity (*<* 1.5 cm) was modestly decreased, while it was unchanged farther away. Increasing the myocardial CV increased the magnitude of the CV error, approximately by the same factor as the increase in CV.

## 4 Discussion

This study performed a set of simulations to ascertain the accuracy of using surface-only activation measurements to determine the CV, and assess tissue health. Several recording resolutions were examined, as well as the effects of the Purkinje system, tissue structure, and relative CV. The major findings are that 1) CV can be measured accurately within 2 cm of the pacing site, 2) spatial sampling of at most 1 mm is necessary, and 3) tissue structure and the Purkinje system were equally responsible for CV measurements far from the pacing site.

### 4.1 Accuracy

The CV measured near the site of pacing was the most accurate. Accepting a maximum error of 10 cm/s, about 10% of the longitudinal CV, CV measured within 2 cm of the pacing site was acceptable and the CV increased rapidly beyond that. CV measured in the nonstimulated ventricle was not reliable.

There are several reasons for the discrepancy between true CV and surface measured CV. The inaccuracy of capturing behaviour which is fundamentally 3D from surface recordings has been shown [3, 4]. This is especially apparent on the septal region of the unactivated ventricular endocardium, where large breakthrough occur as activity propagates across the septum.

The orientation of the myocytes has a large effect on the surface propagation, roughly equal to that of the PS. Removing both of these aspects results in a propagation that is orthogonal to the surface and is accurately measured by surface-only measurements, with error decreased by an order of magnitude. This study highlights how interpreting surface activation patterns over large regions as simple planar wave propagation in isotropic tissue leads to misleading conclusions. Propagation is complicated and has to be understood in the context of complex structure architecture with fast auxiliary pathways.

### 4.2 Purkinje System

The Purkinje system is also a factor leading to surface CV error. The electrical activity propagates into the PS and then exits farther away from the pacing site. The PS activity can breakthrough ahead of the myocardial wavefront, or merge with it, and effectively speed up the apparent prop-agation. This situation between PS and myocardial interplay is further complicated by delays entering and exiting the PS, with anterograde delays being longer. Near the pacing site, these delays are longer than the time to propagate through the tissue, so the surface propagation is most similar to the myocardial propagation. The activity has also not traversed the septum or collided with any obstacles at this point. Without a PS, the error was much lower (Fig. 5), as would be expected. Also, the initial decrease in CV error followed by a rapid increase in the error was no longer seen without the PS.

### 4.3 Fibrosis

Two different types of fibrosis were examined, dense and diffuse. The PS was kept as fully func-tioning as suggested by several studies [7, 8]. The diffuse fibrotic region was fully transmural but still allowed conduction, albeit greatly slowed in all directions. Regardless of where the ventricles were paced, the diffuse fibrosis area was identifiable as a zone of slow conduction with relatively small CV error. Hence, CV mapping can be used clinically to identify areas of diffuse fibrosis, despite the fact that CV values may not be reliable. Thus, unlike high CVs which are usually spurious, lower CVs are more likely correct. However, although the speed of propagation was slow over the diffuse scar, directionality was not consistent as multiple breakthroughs were present.

Importantly, the dense region of intramural fibrosis was not discernible from CV measurements as there existed a healthy, surviving endocardial layer that masked the infarct. The amplitude of the extracellular potential signals was not considered, but could be taken into account to define this region [12].

### 4.4 Uncertainty in clinical measurements

There are multiple sources of uncertainty in electroanatomic mapping recordings including in the construction of the ventricular anatomy, the movement of the catheter, and the assignment of activation time, particularly in the case of fractionated electrograms. This means that errors in CV measurements due to catheter movement, for example, may be bigger than the errors because of the surface nature of the measurement. Coveney et al developed methodologies for quantifying these uncertainties in CV measurements that could be extended to volumetric measurements [13]. In addition, the methodology used to estimate CV could have an impact on the difference between surface and volumetric estimates. Methodologies that utilise more measurements over a larger area for estimation will be less sensitive to the effects of PS break-through, but may not capture local heterogeneity [14].

### 4.5 Implications for arrhythmia mapping

Identifying the location and conduction properties of reentrant ventricular tachycardia (VT) cir-cuits is important for targeting catheter ablation therapy [15]. Our study indicates the importance of the three-dimensional nature of the ventricular tissue and its conduction, which should be con-sidered when mapping VT circuits. Future work will simulate different circuits and compare their surface and volumetric CV estimates and propagation vector field analysis.

The travelling isotropic planar wave is the simplest propagation pattern to conceive and analyze. If this were the case, the apparent CV would be accurate everywhere, as shown in Fig. 6. Recently, a hyperboloid form has been suggested as an excitation pattern for monomorphic ventricular circuits [16]. However, even this more sophisticated activation pattern interprets surface isochrones assuming isotropic propagation with no Purkinje contribution. This surface pattern could, thus, not be truly indicative of the underlying true activation. In contrast to reentry involving a single cylindrical isthmus, histological studies report a more complicated structure with multiple, tortuous routes [17].

**Figure 6.**
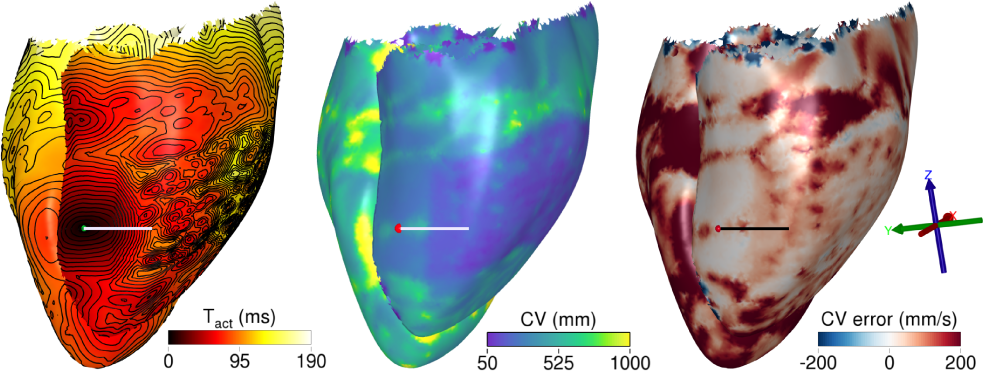
CV and CV error for pacing site #4 near diffuse fibrosis. Right lateral view indicating the RV pacing site by a red dot. **Left**: Activation map with 5 ms isochrones. **Middle**: Conduction velocity (CV) calculated on the endocardial surfaces of the heart. The white scale bar indicates a distance of 2 cm. **Right**: The CV error on the endocardial surfaces is shown with a black scale bar of length 2 cm. The map was computed with 1 mm spacing.

**Figure 7.**
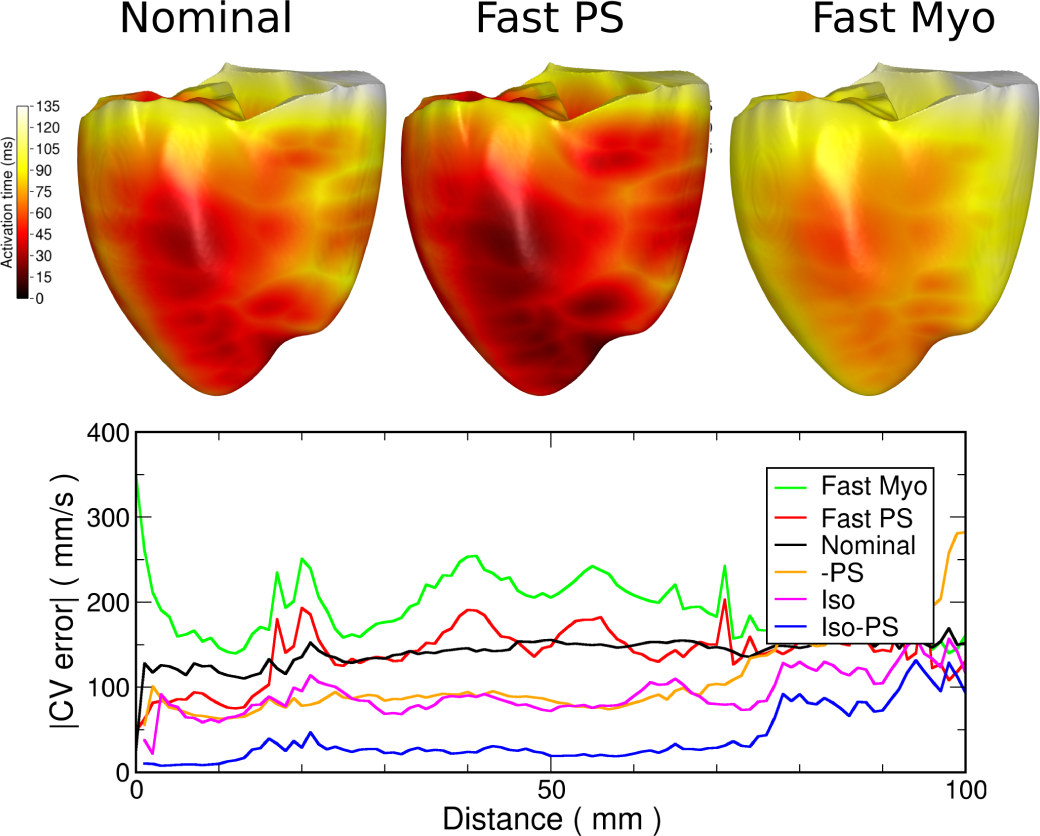
Effect of relative conduction velocities. **Top**: Activation times for the nominal case, Purkinje system CV increased by 1.41 fold (Fast PS), and myocardial CV increased by 1.41 fold (Fast Myo). **Bottom**: Magnitude of conduction velocity error as a function of distance from the pacing site when increasing propagation CV in the myocardium 1.41-fold (Fast Myo), increasing CV in the Purkinje System by 1.41-fold (Fast PS), removing the PS (-PS), ignoring fibre structure (Iso), and ignoring both the PS and the fibre structure (Iso-PS).

A better method for estimating true CV would be to pace and map from the same catheter, moving the catheter over the endocardial surface and stitching together the resultant CVs. The catheter should ideally been high density and have an area of several cm^2^. Measuring CV in the same ventricle as the one paced, as our study suggests, has been previously promoted to identify poorly coupled myocardium [16].

### 4.6 Conclusions

Conduction velocity can be accurately measured by surface electrodes if the distance between recording electrodes is at most 1 mm, and if the recordings are made in the range of 0.5–2 cm from the stimulating site. Diffuse transmural fibrosis is detectable, irrespective of distance from the pacing site.

### 4.7 Limitations

Sinus activation of the ventricles, approximating the QRS width was longer than normal, being 135 ms instead of a more normal value of 120 ms. This had little bearing on the results as shown in §3.6.

## 5 Sources of Funding

EJV and JB were supported by the French Government as part of the “Investments of the Future” program managed by the National Research Agency (ANR), Grant reference ANR-10-IAHU-04. JB was also supported by the European Union’s Horizon 2020 research and innovation program under the ERA-NET co-fund action No. 680969 with ANR [ERA-CVD SICVALVES grant ANR-19-ECVD-0006]. CR acknowledges support from a UKRI Future Leaders Fellowship (MR/W004720/1). This work was performed using HPC resources from GENCI–IDRIS (Grant A0140310517).

## Data Availability

These are computer simulations with no clinical data.

## Notes

### Competing Interest Statement

The authors have declared no competing interest.

### Clinical Trial

N/A

### Funding Statement

EJV and JB were supported by the French Government as part of the ?Investments of the 267 Future? program managed by the National Research Agency (ANR), Grant reference ANR-10-268 IAHU-04. JB was also supported by the European Union?s Horizon 2020 research and innovation 269 program under the ERA-NET co-fund action No. 680969 with ANR [ERA-CVD SICVALVES 270 grant ANR-19-ECVD-0006]. CR acknowledges support from a UKRI Future Leaders Fellowship 271 (MR/W004720/1). This work was performed using HPC resources from GENCI?IDRIS (Grant 272 A0140310517).

## References

1. Boyle PM, Franceschi WH, Constantin M, Hawks C, Desplantez T, Trayanova NA, Vigmond EJ. New Insights on the Cardiac Safety Factor: Unraveling the Relationship Between Conduction Velocity and Robustness of Propagation. Journal of Molecular and Cellular Cardiology, 2019;128:117–128.

2. Takahashi Y, Yamaguchi T, Otsubo T, Nakashima K, Shinzato K, Osako R, Shichida S, Kawano Y, Fukui A, Kawaguchi A, et al. Histological Validation of Atrial Structural Re-modelling in Patients with Atrial Fibrillation. European Heart Journal, 2023;ehad396.

3. Padilla JR, Anderson RD, Joens C, Masse S, Bhaskaran A, Niri A, Lai P, Azam MA, Lee G, Vigmond E, Nanthakumar K. Orientation of Conduction Velocity Vectors on Cardiac Mapping Surfaces. *Europace : European Pacing, Arrhythmias, and Cardiac Electrophysiology: Journal of the Working Groups on Cardiac Pacing*, Arrhythmias, and Cardiac Cellular Electrophysiology of the European Society of Cardiology, 2023;25:1172–1182.

4. Anderson RD, Rodriguez Padilla J, Joens C, Masse S, Bhaskaran A, Magtibay K, Niri A, Asta J, Lai P, Azam MA, Vigmond E, Nanthakumar K. On the Electrophysiology and Mapping of Intramural Arrhythmic Focus. Circulation. Arrhythmia and Electrophysiology, 2022;15:e010384.

5. Bayer JD, Sobota V, Moreno A, Jäıs P, Vigmond EJ. The Purkinje Network Plays a Major Role in Low-Energy Ventricular Defibrillation. Computers in Biology and Medicine, 2022; 141:105133.

6. Bayer J, Prassl AJ, Pashaei A, Gomez JF, Frontera A, Neic A, Plank G, Vigmond EJ. Universal Ventricular Coordinates: A Generic Framework for Describing Position within the Heart and Transferring Data. Medical Image Analysis, 2018;45:83–93.

7. Friedman PL, Fenoglio JJ, Wit AL. Time Course for Reversal of Electrophysiological and Ultrastructural Abnormalities in Subendocardial Purkinje Fibers Surviving Extensive My-ocardial Infarction in Dogs. Circulation Research, 1975;36:127–144.

8. Friedman PL, Stewart JR, Fenoglio JJ, Wit AL. Survival of Subendocardial Purkinje Fibers After Extensive Myocardial Infarction in Dogs. Circulation Research, 1973;33:597–611.

9. Vigmond EJ, Hughes M, Plank G, Leon LJ. Computational Tools for Modeling Electrical Activity in Cardiac Tissue. Journal of Electrocardiology, 2003;36 Suppl:69–74.

10. Roth BJ. Electrical Conductivity Values Used with the Bidomain Model of Cardiac Tissue. IEEE Transactions on Bio-Medical Engineering, 1997;44:326–328.

11. Glukhov AV, Fedorov VV, Kalish PW, Ravikumar VK, Lou Q, Janks D, Schuessler RB, Moazami N, Efimov IR. Conduction Remodeling in Human End-Stage Nonischemic Left Ventricular Cardiomyopathy. Circulation, 2012;125:1835–1847.

12. Soejima K, Stevenson WG, Maisel WH, Sapp JL, Epstein LM. Electrically Unexcitable Scar Mapping Based on Pacing Threshold for Identification of the Reentry Circuit Isthmus: Feasibility for Guiding Ventricular Tachycardia Ablation. Circulation, 2002;106:1678–1683.

13. Coveney S, Corrado C, Roney CH, O’Hare D, Williams SE, O’Neill MD, Niederer SA, Clay-ton RH, Oakley JE, Wilkinson RD. Gaussian Process Manifold Interpolation for Probabilis-tic Atrial Activation Maps and Uncertain Conduction Velocity. Philosophical Transactions of the Royal Society A, 2020;378:20190345.

14. Coveney S, Cantwell C, Roney C. Atrial Conduction Velocity Mapping: Clinical Tools, Algorithms and Approaches for Understanding the Arrhythmogenic Substrate. Medical & Biological Engineering & Computing, 2022;60:2463–2478.

15. Ciaccio EJ, Anter E, Coromilas J, Wan EY, Yarmohammadi H, Wit AL, Peters NS, Garan H. Structure and Function of the Ventricular Tachycardia Isthmus. Heart Rhythm, 2022; 19:137–153.

16. Nishimura T, Shatz N, Weiss JP, Zawaneh M, Bai R, Beaser AD, Upadhyay GA, Aziz ZA, Nayak HM, Shatz DY, Miyazaki S, Goya M, Sasano T, Su W, Raiman M, Tung R. Identification of Human Ventricular Tachycardia Demarcated By Fixed Lines of Conduction Block in a 3-Dimensional Hyperboloid Circuit. Circulation, 2023;148:1354–1367.

17. Glashan CA, Androulakis AFA, Tao Q, Glashan RN, Wisse LJ, Ebert M, de Ruiter MC, van Meer BJ, Brouwer C, Dekkers OM, Pijnappels DA, de Bakker JMT, de Riva M, Piers SRD, Zeppenfeld K. Whole Human Heart Histology to Validate Electroanatomical Voltage Mapping in Patients with Non-Ischaemic Cardiomyopathy and Ventricular Tachycardia. European Heart Journal, 2018;39:2867–2875.

